# Reconsidering Silent Variant Unveils *SGCA*’s Role in Atypical Cardiomyopathy

**DOI:** 10.1101/2025.02.10.25320701

**Authors:** Smadar Horowitz-Cederboim, Ronit Hoffman-Lipschuetz, Ronen Durst, Tamar Harel, Shoshi Shpitzen, Ayelet Shauer, Donna R Zwas, Avital Eilat, Orr Tomer, Vardiella Meiner

## Abstract

We investigated an atypical form of limb-girdle muscular dystrophy type R3 (LGMDR3) associated with a homozygous synonymous variant in *SGCA* that significantly expands the recognized clinical phenotype to include prominent cardiac involvement. While LGMDR3, caused by pathogenic variants in *SGCA*, typically presents as progressive muscle weakness with relatively limited cardiac manifestations, the individuals in our study developed substantial left ventricular dysfunction, arrhythmias, and even life-threatening ventricular tachyarrhythmias.

Five consanguineous families, initially evaluated through exome sequencing, showed no conclusive results until a focused genotype-phenotype correlation reanalysis pinpointed a single homozygous synonymous *SGCA* variant. Despite its location distant from canonical splice sites, this variant disrupted normal mRNA splicing, leading to aberrant transcripts and, presumably, a nonfunctional or structurally altered α-sarcoglycan protein. Cardiac assessments, including echocardiography, Holter monitoring, and cardiac magnetic resonance imaging, revealed a spectrum of findings from mild asymptomatic dysfunction to severe dilated cardiomyopathy and malignant arrhythmias requiring implantable cardioverter-defibrillators.

This case broadens current understanding of LGMDR3 and its molecular underpinnings, ultimately guiding more personalized management strategies for affected individuals. It illustrates that thorough and ongoing variant interpretation is crucial, particularly in consanguineous populations where seemingly benign alterations may confer significant phenotypic consequences. By integrating carefully curated genomic data with clinical insights, clinicians can better identify patients at risk, tailor surveillance protocols, and implement timely interventions. In doing so, this discovery not only refines our appreciation of the intricate interplay between genetic variants and their phenotypic manifestations, but also underscores the importance of precision medicine approaches in the evolving landscape of inherited cardiomyopathies.

## Introduction

The genetic landscape of inherited cardiomyopathies is characterized by a complex array of inheritance patterns. While autosomal dominant transmission is most common in monogenic forms, the field has increasingly recognized the importance of autosomal recessive, X-linked, and mitochondrial inheritance modes ^1^. Recessive cardiomyopathies are significant in consanguineous populations and may co-occur with additional features observed in myopathy phenotypes.

Limb-girdle muscular dystrophy type R3 (LGMDR3), previously known as LGMD2D ^2^, is caused by pathogenic variants in the *SGCA* gene. It is characterized by progressive proximal muscle weakness, with clinical severity ranging from asymptomatic cases to severe early-onset forms. Unlike other muscular dystrophies, cardiac involvement in LGMDR3 is relatively uncommon. Studies suggest that overt cardiomyopathy is observed in approximately 10% of LGMDR3 patients ^3,4^.

Here, we describe findings from five consanguineous families in which affected individuals had undergone exome testing for various indications, including cardiac symptoms with muscle involvement, isolated myopathy, or both. Initial analyses provided no conclusive results. However, subsequent reanalysis incorporating genotype-phenotype correlation within our cohort revealed a homozygous synonymous variant in *SGCA*. This variant led to an atypical form of LGMDR3 characterized by significant cardiac involvement.

This discovery highlights the critical value of periodic genetic data reanalysis, especially when confronted with complex and seemingly unrelated clinical presentations across families. It also underscores the need for careful evaluation of variants initially classified as benign, given their potential to hold substantial clinical significance.

## Methods

### Ethical considerations

All participants provided written informed consent for clinical data collection and genetic studies, approved by the Hadassah Medical Center Helsinki committee (0151-20-HMO).

### Exome sequencing

Genomic DNA was extracted from whole blood samples of the following individuals: A-II-1, A-II-3, B-III-1, C-III-1; D-III-2; E-II-1 and E-II-3. Exonic sequences from DNA were enriched with the SureSelect Human All Exon 50 Mb V5 Kit (Agilent Technologies, Santa Clara, California, USA), or with the xGen Exome Research Panel IDT-V2 combined with the xGen Human mtDNA Research Panel v1.0 (IDT). Sequences were generated on a HiSeq2500 or NovaSeq6000 sequencing system (Illumina, San Diego, California, USA) as 125- or 150-bp paired-end runs. Read alignment and variant calling of single nucleotide variants, structural variants and copy number variants were done on the Geneyx Analysis platform^5^ using Illumina DRAGEN Bio-IT. The human genome assembly hg19 (GRCh37) was used as reference. Variants were annotated on the Geneyx Analysis annotation engine. Exome analysis of the probands yielded 44-105 million reads, with a mean coverage of 63-136X.

### Segregation analysis

An amplicon containing the *SGCA* variant was amplified by conventional PCR of genomic DNA from probands and all available parents and siblings and analyzed by Sanger dideoxy nucleotide sequencing with forward and reverse primers as indicated in **Table S1.**

### Clinical evaluation

All individuals homozygous for the variant underwent comprehensive evaluation, including blood tests for hepatocellular enzymes and creatine phospho-kinase (CPK) levels, a resting electrocardiogram (ECG), 24-hour Holter monitoring 3 leads (Lifecard, Spacelabs Healthcare, Snoqualmie, Washington, United States), two-dimensional echocardiography, and cardiac magnetic resonance (CMR) imaging. Participants were scanned on either 1.5T Avanto Siemens (Munich, Germany) or 3T Philips Ingenia (Amsterdam, Netherlands) scanners. Assessment of left and right ventricular (LV and RV, respectively) systolic function on CMR was performed in accordance with recent criteria for the evaluation of cardiomyopathies. Late gadolinium enhancement (LGE) imaging was performed 10-15 minutes after intravenous administration of 0.2 mmol/kg of gadoterate-meglumine (Dotarem, manufactured by Guerbet, Villepinte, France) via short-axis breath-hold phase-sensitive inversion recovery (PSIR) images scanning the entire length of the myocardium. The endocardial and epicardial borders of the LV myocardium in end-diastole and end-systole were traced manually on the short-axis slices. The percent delayed enhancement was automatically determined as 5 SD above a sample of normal remote myocardium as defined by the reader (RD). The analysis was done on 3D Synapse software by Fuji. After completing the evaluation, each patient was seen in consultation by a cardiologist specializing in inherited arrhythmias.

### Cell culture and generation of cell lines

Primary fibroblasts were grown from skin-punch biopsies and maintained in Dulbecco’s modified Eagle’s medium (DMEM) supplemented with 15% FCS, 1% L-glutamine, and 1% penicillin-streptomycin antibiotics (Biological Industries Beit Haemek, Israel). Primary fibroblasts were immortalized with a lentivirus expressing human telomerase reverse transcriptase (hTERT) that was generated by transfecting HEK293T cells (ATCC) with the plasmids pLV-UBC-hTERT (Addgene no. 114316), psPax2 (Addgene no. 12260), and pMD2.G (Addgene no. 12259). The selection was performed using blasticidin (Bio prep) at 10 μg/mL.

### RNA/cDNA analysis

RNA was isolated from patient (B-III-1) and control fibroblasts by TRIzol reagent extraction. cDNA was prepared from 1 μg RNA using the qScript cDNA Synthesis Kit (Quantabio). Target regions within *SGCA* were amplified by PCR reactions using VeriFi™ DNA Polymerase Mix Red (PCRBIOSSTEMS) with forward and reverse primers as indicated in **Table S1**. Housekeeping gene *B-actin* served as loading controls to normalize expression levels. The resultant fragments were separated by 1.5% (w/v) agarose gel electrophoresis, and their sequences were determined by Sanger sequencing.

## Results

### Clinical Presentation

We present three representative cases, all males aged 20-30 years at their diverse first major clinical presentation.

#### Patient A-II-1

A male aged 21-25 years presented with sudden palpitations, chest pain, and weakness during physical activity. His medical history included elevated creatine kinase (CK) levels and episodes of rhabdomyolysis since early childhood, similar to other affected individuals in the same family. He was diagnosed with non-ischemic dilated cardiomyopathy (NIDCM). CMR showed LGE in the lateral wall at the subepicardial and mid-myocardial layers. He underwent placement of an implantable cardioverter-defibrillator (ICD) due to recurrent polymorphic ventricular tachycardia. Despite medical treatment, he experienced recurrent appropriate ICD shocks, requiring multiple ablation procedures, including an epicardial approach. Ventricular arrhythmias have been well controlled over the past four years.

#### Patient A-II-3

A male aged 26-30 years presented with myocarditis-like during early adulthood. His medical history included elevated CK levels since childhood and hepatitis C carrier status. Initially, his cardiac function was normal but later declined to biventricular dysfunction (LVEF 40%, RVEF 42%). CMR revealed LGE in the lateral and inferior walls, with no further progression. He has not experienced major ventricular arrhythmias to date, although a 6-beat monomorphic NSVT was recorded on Holter monitoring. Due to complaints about palpitation episodes, an internal loop recorder (ILR) was implanted. Treatment with ACE inhibitors and beta-blockers restored his ventricular function to normal after several months of therapy.

#### Patient B-III-1

A male aged 26-30 years was successfully resuscitated following ventricular fibrillation (VF) during physical activity. His medical history included elevated creatine kinase (CK) levels and untreated cardiomyopathy first identified during adolescence, with a left ventricular ejection fraction (LVEF) of 45%. Another male in the same family died suddenly in late adolescence. Echocardiography revealed severely decreased systolic function (LVEF <20%) with left ventricular dilatation. CMR demonstrated LGE in the mid-myocardial layers of the basal inferior and lateral walls. Holter monitoring showed frequent premature ventricular contractions (PVCs), up to 23%, mostly parasystolic beats originating from a left fascicle. These PVCs were refractory to antiarrhythmic medications. He underwent ICD implantation and later an ablation of those PVCs. Although successfully ablated, no improvement in systolic function was observed.

Overall, 13 individuals are reported in this study. Clinical manifestations are summarized in **Table 1**. The mean age at first presentation (elevated CK levels in 8/11, myopathy in 3/11) was within the range of 6-18 years, while the mean age at the last clinical evaluation was within the range of 12-39 years, with 50% being female. All tested individuals (11/11, 100%) demonstrated elevated CK levels and elevated hepatocellular enzymes (ALT, AST at 2–3× the upper normal limit). Additionally, 55% (6/11) reported progressive muscle weakness or myalgia, while 45% (5/11) experienced recurrent episodes of rhabdomyolysis. Notably, heterozygous carriers did not exhibit any clinical signs or symptoms to date.

**Table 1.**
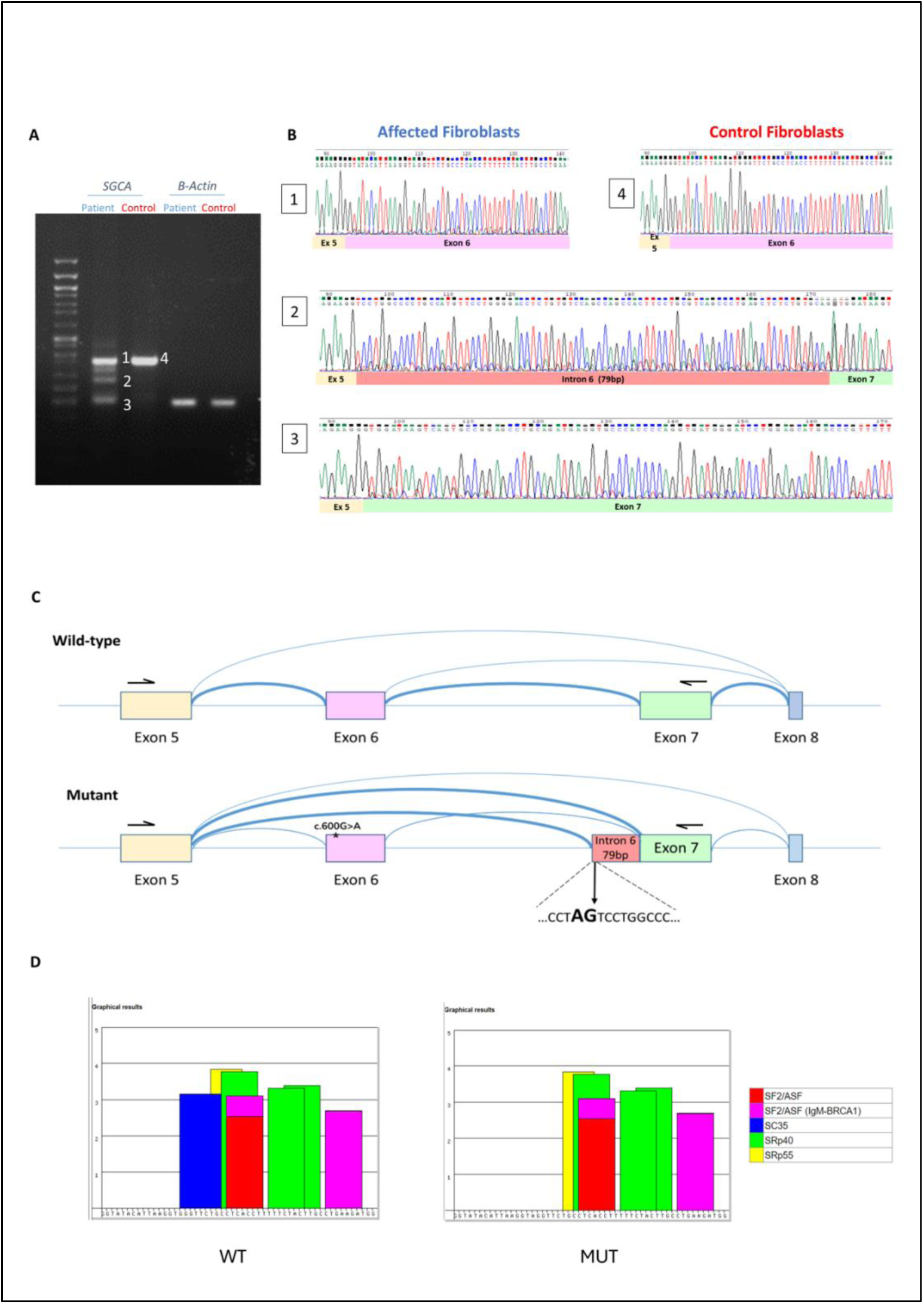
Clinical characteristics.

Among 12 evaluated subjects, 50% (6/12) exhibited a decline in systolic LV function, ranging from mild asymptomatic dysfunction to severe impairment. CMR revealed abnormal findings in five individuals (100% of tested individuals, 5/5), primarily LGE in the lateral (5/5) and inferior (4/5) walls, localized to the subepicardial and mid-myocardial layers (**Figure S1B, C**). LV dilatation was observed in two individuals, while the remaining four were classified as having non-dilated left ventricular cardiomyopathy (NDLVC). ECG abnormalities were observed in 50% (3/6), including low voltage in limb leads, incomplete right bundle branch block (IRBBB), and other findings.

Two subjects experienced multiple episodes of polymorphic ventricular tachycardia/ventricular fibrillation (PMVT/VF), necessitating ICD placement. Another subject demonstrated a short monomorphic NSVT on Holter monitoring. Two additional subjects exhibited a burden of PVCs, not exceeding 1,000 beats per 24 hours. In family B, individual III-6 died suddenly in late adolescence (16-20 years) from unknown causes, precluding genetic testing due to the unavailability of DNA.

### Exome sequencing reveals a shared homozygous variant in SGCA

Whole-exome sequencing was undertaken on 6 individuals from 4 seemingly unrelated families. Initial analysis was uninformative. Subsequent reanalysis, aided by genotype and phenotype matching within our dataset of 22,400 exomes with various referral indications, identified a homozygous synonymous variant in the *SGCA* gene [MIM: 600119] – chr17:50169107G>A [hg38]; NM_000023.4: c.600G>A p.Val200= in all affected individuals. All families were from a geographically isolated consanguineous population. The two brothers in Family A shared a region of homozygosity (ROH) around the variant, as did the other three individuals, suggesting a founder variant. The *SGCA* variant was located near the boundaries of both ROH regions, such that the maximum overlap between all individuals was only 0.65MB. The allele frequency was 0.07% in our dataset and elevated within this population. Following identification of this variant, three additional families with overlapping phenotypes were found to harbor the same variant.

The *SGCA* variant was initially overlooked due to its synonymous nature and distance from canonical splice sites, i.e. located 15 bp from the nearest exon boundary. However, the recurrence of the variant within the database and the overlapping phenotype between the cases led to further evaluation of the variant.

#### SGCA variant leads to aberrant splicing

The SpliceAI algorithm ^6^ predicted that the *SGCA* variant may alter splicing (splice-altering moderate, 0.31). Furthermore, the variant was predicted to abolish an exonic splicing enhancer (ESE) motif recognized by the serine/arginine-rich (SR) splicing factor, SRSF2 (also known as SC35) (**Figure 2D)** ^7,8^. To establish the effect of the variant at the RNA level, we analyzed the cDNA sequence derived from fibroblasts of an affected individual vs. control. RT-PCR revealed amplicons that did not appear in the control (**Figure 2A**). Sanger sequencing demonstrated two primary aberrant transcripts: one skipping exon 6 entirely (149 bp), and a second transcript with an intronic inclusion of 79 bp from intron 6 adjacent to exon 7 (**Figure 2B**-**C**). The 79 bp sequence from intron 6 was preceded by a consensus acceptor splice site (**Figure S2**). Presumably, loss of the ESE led to skipping of exon 6 and recognition of an alternative splice acceptor site within intron 6. To ensure that there was no other intronic sequence variant in intron 6 which led to the mis-splicing, we sequenced intron 6 by the Sanger method. No other variant was identified, underscoring the pathogenicity of the synonymous c.600G>A p.Val200= variant.

**Figure 1.** Pedigrees of five families harboring the *SGCA* variant. Figure 1 has been removed to protect patient confidentiality. Readers interested in accessing this information may contact the corresponding author upon reasonable request

**Figure 2.**
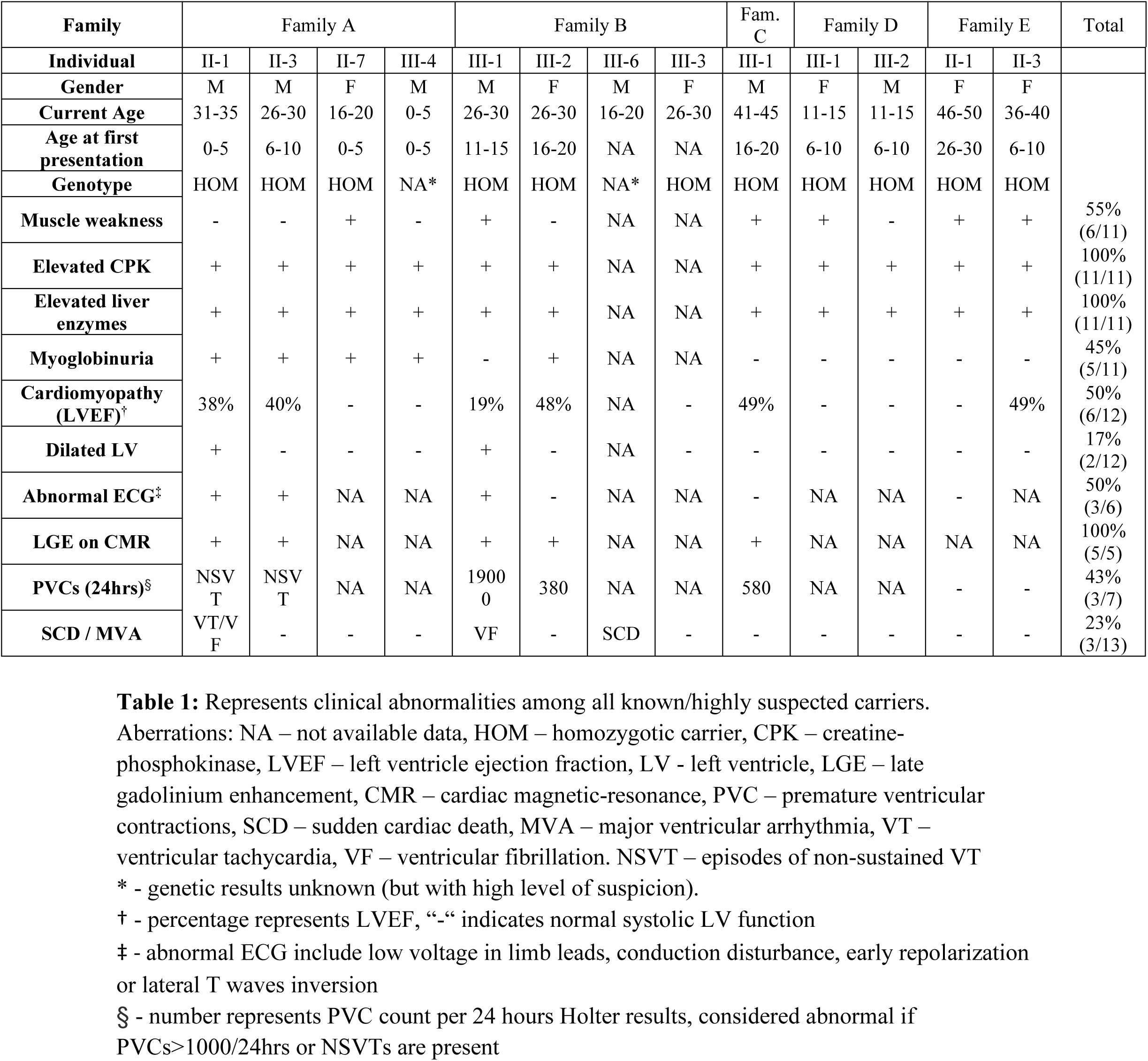
cDNA analysis of *SGCA* variant indicates aberrant splicing. (A) Gel electrophoresis indicating aberrant splicing in affected individual vs. control. Four bands were cut from the gel and subject to Sanger sequencing. *B-actin* was used as a housekeeping gene. (B) Sanger sequencing of cDNA from fibroblasts of affected individual (left panels) and wild-type control (right panel). (C) Diagram of the exon skipping and intronic inclusion. (D) The *SGCA*:c.600G>A variant cancels the exonic splicing enhancer motif recognized by SC35, according to the ESE Finder database prediction (https://esefinder.ahc.umn.edu/cgi-bin/tools/ESE3/esefinder.cgi?process=home).

## Discussion

### Expanded Phenotype of LGMDR3 with Cardiac Involvement

This study establishes a connection between a synonymous *SGCA* variant and an expanded LGMDR3 phenotype that prominently includes significant cardiac involvement. Traditionally, *SGCA* variants have been associated with LGMDR3, characterized by a variable phenotype including proximal muscle weakness, elevated CPK levels, calf muscle pseudohypertrophy, and mobility challenges. Notably, all subjects in this study displayed elevated liver enzymes, a feature previously documented in other LGMD subtypes ^9,10^.

The findings in this cohort extend the recognized clinical spectrum of LGMDR3, revealing a substantial incidence of cardiac involvement. Specifically, 50% of the tested individuals (6 out of 12) exhibited varying degrees of left ventricular dysfunction, ranging from mild, asymptomatic impairment to severe dysfunction. Additionally, several individuals experienced life-threatening ventricular arrhythmias, including two cases of ventricular tachycardia/fibrillation (VT/VF), one instance of non-sustained ventricular tachycardia (NSVT), and two cases of mild PVCs. Tragically, one family member suffered sudden cardiac death (SCD), although his genotype could not be determined.

The unusual cardiac involvement observed in our cohort is particularly noteworthy when compared to previous reports on LGMDR3. Pathogenic variants in *SGCA* are typically associated with low rates of cardiomyopathy, primarily in cases involving exon 3 variants ^4^. A comparative study indicated that cardiomyopathy and cardiac events are less frequent in LGMDR3 than in LGMDR5, with only 12% of LGMDR3 patients exhibiting left ventricular dysfunction. The mean LVEF in LGMDR3 patients was reported as 63%, although an LVEF below 55% was correlated with increased mortality ^11^.

It is important to highlight, however, that the existing literature on cardiac involvement in LGMDR3 is based on small cohort studies, emphasizing the need for more extensive research in this area. In the broader context of limb-girdle muscular dystrophies (LGMD), encompassing both autosomal dominant and recessive forms, LGE is a commonly observed finding in CMR imaging of patients with LV dysfunction. LGE typically affects less than 10% of LV mass, predominantly appearing in the subepicardial layer or as patchy infiltrates in the mid-myocardial layer, particularly in the lateral wall ^12^. Unlike other cardiomyopathies, the prognostic significance of LGE in LGMD remains undefined.

### Molecular Mechanisms and Functional Impact of the SGCA Variant

To better understand the molecular basis of these findings, it is essential to examine the role of *SGCA.* This gene encodes α-sarcoglycan, a key component of the sarcoglycan complex within the larger dystrophin-associated protein complex (DAPC) in skeletal and cardiac muscles. The DAPC plays an essential role in preserving sarcolemmal integrity during the cycles of muscle contraction and relaxation. Pathogenic *SGCA* variants include missense, nonsense, frameshift, and splice site mutations ^3,4^.

Interestingly, the synonymous variant *SGCA:* c.600G>A caused significant alterations in mRNA splicing, despite neither altering the amino acid sequence nor being near a canonical splice site. RNA/cDNA analysis demonstrated that this variant, located in exon 6 (of 10), likely disrupts an ESE domain, which is critical for spliceosome positioning ^13^. The variant was predicted to abolish the ESE motif recognized by the SR splicing factor SRSF2/SC35, thereby promoting exon 6 skipping during the splicing process. This disruption results in two primary aberrant transcripts: one that skips exon 6 *(5-7-8)* and another that skips exon 6 while incorporating 79 nucleotides from intron 6 adjacent to exon 7 *(5-intron6partial-7-8)*. The exon-skipping transcript introduces a frameshift, potentially leading to a truncated or nonfunctional protein. Interestingly, the inclusion of 79 intronic nucleotides compensates for the frameshift caused by exon 6 skipping, preserving the reading frame. However, this transcript remains abnormal, as it lacks exon 6 and includes an intronic sequence. The functional consequences of this altered mRNA structure are yet to be determined and warrant further investigation.

### Clinical Management and Implications for Risk Stratification

These findings carry significant implications for clinical management. According to the 2022 Heart Rhythm Society expert consensus statement on arrhythmic risk management in neuromuscular disorders ^14^, ICD placement is recommended for primary prevention of ventricular arrhythmias in LGMD2 (recessive forms of LGMD) patients with an LVEF below 35%. However, based on the arrhythmic history observed in the families studied here, shared decision-making discussions were conducted with patients exhibiting any degree of LV dysfunction and LGE on CMR. This personalized approach may be particularly critical for individuals with this specific *SGCA* variant, given its association with marked cardiac involvement.

The clinical implications of our findings are profound, particularly in light of the observed cardiac manifestations, including cases of sudden cardiac death. Our internal database indicates an allele frequency of 5.1% for this *SGCA* variant within the village population. Due to the high rates of consanguinity, accurately estimating the carrier rate remains challenging. To address this significant health risk, we propose the implementation of a comprehensive population screening strategy. Such an approach aims to identify at-risk individuals early, thereby transforming preventive care and management strategies for affected families. The efficacy of early intervention is demonstrated by index patient 2 (A-II-3), whose ventricular function improved following targeted medical care.

### Re-evaluating Variant Classification

This case underscores the importance of reevaluating existing variant classifications especially in consanguineous populations where rare variants are more prevalent. The identification of this significant synonymous *SGCA* variant, initially overlooked, reveals the complexities of variant interpretation and emphasizes the necessity of integrating clinical expertise with iterative genetic analysis. Reevaluating variant classifications considering new clinical evidence can lead to more accurate diagnoses and improved patient care.

This finding also illustrates the limitations of relying solely on database annotations, as evidenced by the variant’s prior classification as likely benign (LB) in ClinVar ^15^. While resources like ClinVar provide valuable platforms for unifying knowledge across laboratories, clinical geneticists should prioritize thorough analysis of the specific family under investigation. By extracting maximum information from internal data before consulting external sources, a more context-specific and comprehensive interpretation of genetic variants can be achieved. Importantly, the global description of this variant in ClinVar underscores its broader significance beyond our studied population, reinforcing the need for continuous reevaluation of genetic variants considering emerging clinical and functional data across diverse populations.

## Limitations and Future Directions

Our study presents several noteworthy limitations. Firstly, RNA expression analysis was conducted in fibroblasts, a suboptimal tissue choice for this condition, due to lack of expression in blood. Ideally, cardiac and skeletal muscle samples would provide more relevant data for the observed phenotype, yet were not available. Secondly, our study is limited by a small sample size drawn from a single village, potentially affecting the generalizability of our findings. Moreover, while index cases underwent more comprehensive assessments, the overall small cohort size may introduce bias in our clinical estimations.

These limitations point to several avenues for future research. Examining the differential expression of *SGCA* in relevant tissues, particularly comparing cardiac and skeletal muscle samples, would provide valuable insights into the tissue-specific effects of this variant. Exploring potential therapeutic approaches, including gene therapy and base-editing techniques, could pave the way for targeted treatments. Conducting long-term follow-up studies on affected individuals homozygous for this variant would better elucidate the natural history of this variant. Investigating the prevalence and impact of this variant in other populations could reveal its broader significance. Finally, developing improved bioinformatic tools for identifying potentially pathogenic synonymous variants would enhance our ability to detect similar cases in the future.

## Conclusions

Our study identified a homozygous synonymous variant of *SGCA* (NM_000023.4: c.600G>A; p.Val200=) associated with an atypical presentation of LGMDR3, characterized by cardiomyopathy with significant arrhythmia. This finding expands our understanding of the phenotypic spectrum associated with *SGCA* variants and highlights the potential clinical implications of seemingly benign variants. This case underscores several crucial aspects of modern genetic diagnostics: the importance of thorough variant analysis, especially in consanguineous populations; the value of internal population-specific databases; and the need for cautious interpretation of existing variant classifications. This discovery serves as a reminder of the complexity of genotype-phenotype correlations and the need for comprehensive functional studies to fully understand the impact of genetic variations.

Moreover, this discovery exemplifies the evolving landscape of genetic medicine, where each family’s unique genetic profile can enhance our understanding of genetic disorders and inform more precise diagnostic and therapeutic strategies. Continuous reanalysis of genetic data, guided by clinical evidence and functional insights, remains a cornerstone for advancing precision medicine.

## Data Availability

All data produced in the present study are available upon reasonable request to the authors

## Acknowledgements

The authors wish to thank the families for their participation in this study.

## Supplemental Material

Tables S1

Figure S1–S2

## Non-standard Abbreviations and Acronyms

CPK: Creatine Phosphokinase
CMR: Cardiac Magnetic Resonance
DAPC: Dystrophin-Associated Protein Complex
ECG: Electrocardiogram
ESE: Exonic Splicing Enhancer
ILR: Internal Loop Recorder
LGE: Late Gadolinium Enhancement
LGMDR3: Limb-Girdle Muscular Dystrophy Type R3
LV: Left Ventricular
LVEF: Left Ventricular Ejection Fraction
NDLVC: Non-Dilated Left Ventricular Cardiomyopathy
NIDCM: Non-Ischemic Dilated Cardiomyopathy
NSVT: Non-Sustained Ventricular Tachycardia
PMVT: Polymorphic Ventricular Tachycardia
PVCs: Premature Ventricular Contractions
ROH: Region of Homozygosity
RV: Right Ventricular
SCD: Sudden Cardiac Death
SGCA: α-sarcoglycan
VF: Ventricular Fibrillation

## References

1. Arbelo E, Protonotarios A, Gimeno JR, Arbustini E, Arbelo E, Barriales-Villa R, Basso C, Biagini E, Blom NA, De Boer RA, et al. 2023 ESC Guidelines for the management of cardiomyopathies. Eur Heart J [Internet]. 2023 [cited 2024 Jul 10];44:3503–3626. Available from: https://pubmed.ncbi.nlm.nih.gov/37622657/

2. Straub V, Murphy A, Udd B. 229th ENMC international workshop: Limb girdle muscular dystrophies – Nomenclature and reformed classification Naarden, the Netherlands, 17–19 March 2017. In: Neuromuscular Disorders. 2018.

3. Georganopoulou DG, Moisiadis VG, Malik FA, Mohajer A, Dashevsky TM, Wuu ST, Hu CK. A Journey with LGMD: From Protein Abnormalities to Patient Impact. Protein Journal. 2021;40.

4. Carson L, Merrick D. Genotype–phenotype correlations in alpha-sarcoglycanopathy: a systematic review. Ir J Med Sci [Internet]. 2022 [cited 2024 Jul 10];191:2743–2750. Available from: https://link.springer.com/article/10.1007/s11845-021-02855-1

5. Dahary D, Golan Y, Mazor Y, Zelig O, Barshir R, Twik M, Iny Stein T, Rosner G, Kariv R, Chen F, et al. Genome analysis and knowledge-driven variant interpretation with TGex. BMC Med Genomics. 2019;12.

6. Jaganathan K, Kyriazopoulou Panagiotopoulou S, McRae JF, Darbandi SF, Knowles D, Li YI, Kosmicki JA, Arbelaez J, Cui W, Schwartz GB, et al. Predicting Splicing from Primary Sequence with Deep Learning. Cell. 2019;176.

7. Cartegni L, Wang J, Zhu Z, Zhang MQ, Krainer AR. ESEfinder: A web resource to identify exonic splicing enhancers. Nucleic Acids Res. 2003;31:3568–71.

8. Smith PJ, Zhang C, Wang J, Chew SL, Zhang MQ, Krainer AR. An increased specificity score matrix for the prediction of SF2/ASF-specific exonic splicing enhancers. Hum Mol Genet. 2006;15.

9. Lash T, Kraemer RR. Elevated liver enzymes indicating a diagnosis of limb-girdle muscular dystrophy. J Gen Intern Med. 2014;29.

10. Zhu Y, Zhang H, Sun Y, Li Y, Deng L, Wen X, Wang H, Zhang C. Serum enzyme profiles differentiate five types of muscular dystrophy. Dis Markers. 2015;2015.

11. Fayssoil A, Ogna A, Chaffaut C, Chevret S, Guimarães-Costa R, Leturcq F, Wahbi K, Prigent H, Lofaso F, Nardi O, et al. Natural history of cardiac and respiratory involvement, prognosis and predictive factors for long-term survival in adult patients with limb girdle muscular dystrophies type 2C and 2D. PLoS One. 2016;11.

12. Kashyap N, Nikhanj A, Labib D, Prosia E, Rivest S, Flewitt J, Pfeffer G, Bakal JA, Siddiqi ZA, Coulden RA, et al. Prognostic Utility of Cardiovascular Magnetic Resonance– Based Phenotyping in Patients With Muscular Dystrophy. J Am Heart Assoc. 2023;12.

13. Bhat GR, Sethi I, Rah B, Kumar R, Afroze D. Innovative in Silico Approaches for Characterization of Genes and Proteins. Front Genet. 2022;13.

14. Groh WJ, Bhakta D, Tomaselli GF, Aleong RG, Teixeira RA, Amato A, Asirvatham SJ, Cha YM, Corrado D, Duboc D, et al. 2022 HRS expert consensus statement on evaluation and management of arrhythmic risk in neuromuscular disorders. Heart Rhythm. 2022;19.

15. Landrum MJ, Lee JM, Benson M, Brown GR, Chao C, Chitipiralla S, Gu B, Hart J, Hoffman D, Jang W, et al. ClinVar: improving access to variant interpretations and supporting evidence. Nucleic Acids Res. 2018;46:D1062–D1067.

