## Supplementary material for "Reconsidering Silent Variant Unveils *SGCA*’s Role in Atypical Cardiomyopathy": Table S1 Figures S1S2

### *SGCA* Study:

### Supplementary Data

| Gene | Forward Primer | Reverse Primer |
|---|---|---|
| *SGCA* (RNA analysis) | CGCTTCCTCTCAGCCTTGGGG | AGAAGAACGGGTCATGCTCC |
| *SGCA* (Segregation analysis) | AAGTTTCAACAACCCCTGGC | TCACATTGCACCAGTCAACG |
| *SGCA* (Genomic DNA analysis) | ATTGACCAACAGAGCAGGGA | CAGAAGAACGGGTCATGCTC |
| *B-Actin* | GCAGCTCACCATGGATGATG | AGGATGCCTCTCTTGCTCTG |

**Table S1.** Primers for Sanger sequencing and RT PCR at DNA and RNA level.

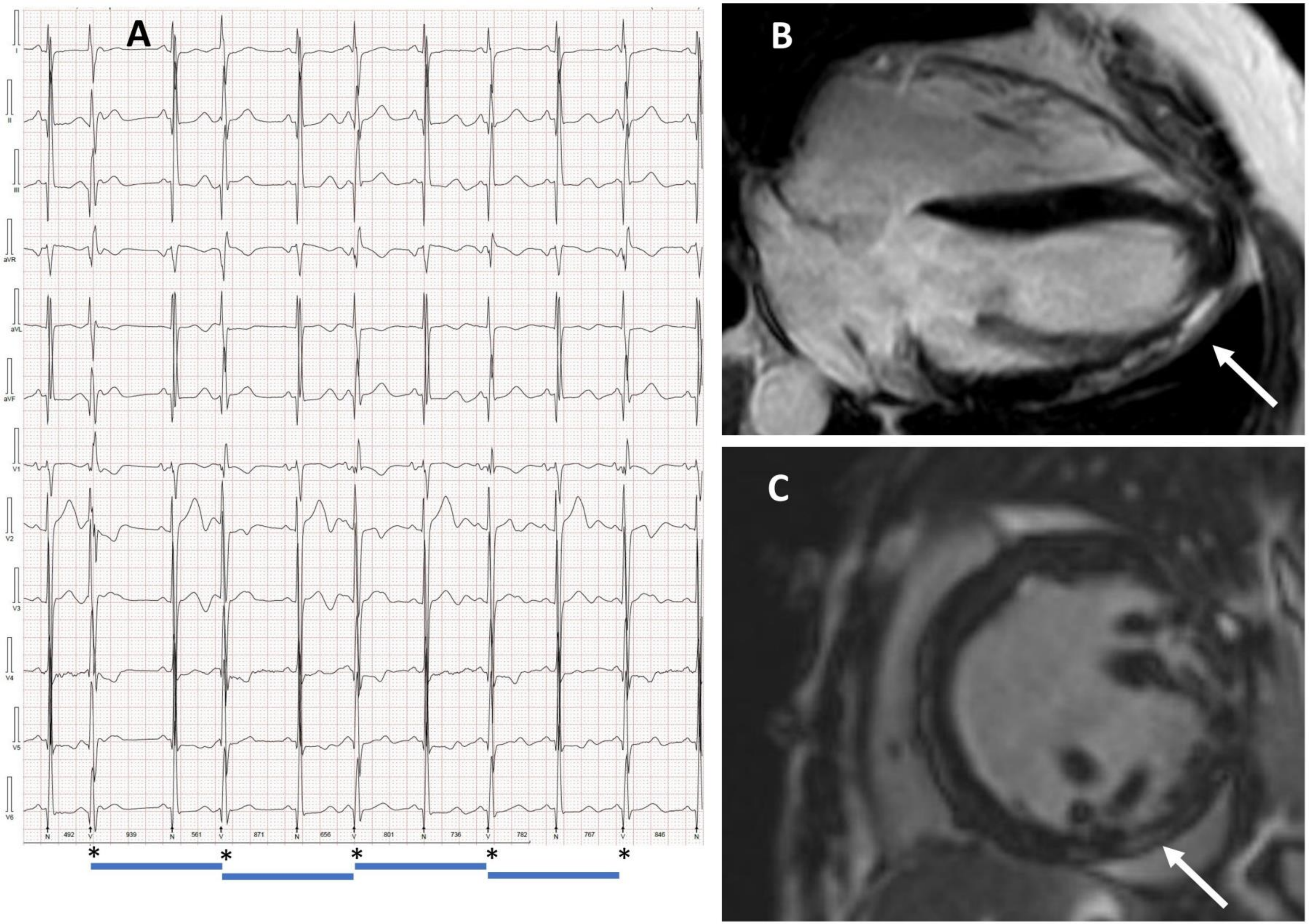


**Figure S1**. **Clinical imaging findings**. (A) Holter recording of subject B-III-1 showing parasystoles originating from the left anterior fascicle. Note the constant intervals between premature beats and varying coupling intervals with sinus beats. (B) Cardiac MRI of subject A-II-3 demonstrating late gadolinium enhancement (LGE) in the inferior wall (white arrow). (C) Cardiac MRI of subject B-III-1 also showing LGE in the inferior wall (white arrow).

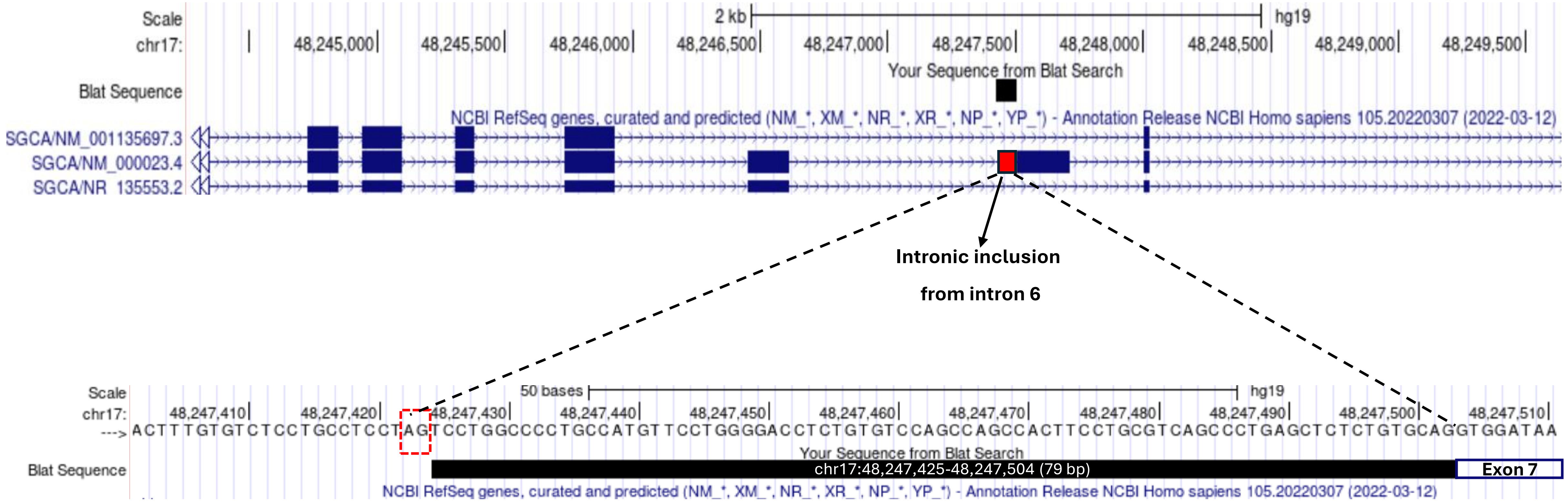


**Figure S2. Sequencing of the SGCA variant at DNA level**. The intronic inclusion (79 bp, red solid box on upper panel) from intron 6 is preceded by a consensus splice acceptor site (AG, red dotted box on lower panel).
